# Unnecessary antibiotic treatment of children hospitalized with RSV-bronchiolitis: risk factors and prescription patterns

**DOI:** 10.1101/2021.07.17.21260687

**Authors:** Uri Obolski, Eias Kassem, Wasef Na’amnih, Shebly Tannous, Viktoria Kagan, Khitam Muhsen

## Abstract

**Background:** Respiratory syncytial virus (RSV) is a main cause of respiratory tract infections, especially affecting young children. Antibiotics are often unnecessarily prescribed for the treatment of RSV. Such treatments can have effects on antibiotic resistance in future bacterial infections of treated patients and the general population.

**Objectives:** We sought to understand the risk factors for and patterns of unnecessary antibiotic prescription in children with RSV.

**Methods:** In a single center retrospective study in Israel, we obtained data of children aged <2 years (n=1015) hospitalized for RSV-bronchiolitis during 2008-2018, and ascertained not to have bacterial coinfections. Antibiotic misuse was defined as prescription of antibiotics during hospitalization of the study population. Patient and clinical variables were assessed as predictors of unnecessary antibiotic treatment in a multivariable logistic regression model.

**Results:** Unnecessary antibiotic treatment rate of children infected with RSV and ascertained not to have a bacterial coinfection was estimated at 33.4% (95% CI 30.5%-36.4%). Increased likelihood of antibiotic misuse was associated with drawing bacterial cultures, and with variables indicative of a severe patient status: lower oxygen saturation, higher body temperature, tachypnea and prior recent emergency room visit. Older age and female sex were also associated with increased likelihood of unnecessary antibiotic treatment.

**Conclusions:** Unnecessary antibiotic treatment in RSV patients was highly common and may be largely attributed to the physicians’ perception of patients’ severity. Improving prescription guidelines, implementing antibiotic stewardship programs and utilizing decision support systems may help achieve a better balance between prescribing and withholding antibiotic treatment.

## Introduction

Respiratory syncytial virus (RSV) is a widespread viral pathogen, causing both upper and lower respiratory tract infections. It is a substantial cause of global morbidity and mortality, especially in young children.^1,2^ RSV typically presents a seasonal pattern, peaking in the winter months in regions of temperate climate, alongside other respiratory viruses.^3^

Accordingly, in Israel RSV is a common cause of respiratory tract infections in young children, though it might cause morbidity in adults as well. ^4,5^ It mainly appears during the winter months (November through March) and is characterized by high attack rate among infants and pre-term born children and comorbidity with pneumonia. ^4–7^

The high healthcare burden attributed to RSV has a secondary deleterious effect - it can increase unnecessary antibiotic usage. Antibiotics are frequently unnecessarily administered to respiratory viral infections, often due to incorrect diagnoses, apprehension from bacterial coinfections, or dismissal of the detrimental effects of unnecessary antibiotic treatment.^8–11^ Accordingly, studies have shown positive temporal associations between incidence of viral respiratory tract infections and antibiotic use. ^12–14^

Redundant antibiotic therapy can be detrimental in two main ways: First, it can harm individual patients by increasing their future likelihood for antibiotic resistant infections^15^ and for antibiotic-associated diarrhea^16^. Second, redundant antibiotic prescription promotes population-wide rates of antibiotic resistance, leading to the dangerously increasing trends of antibiotic resistance observed worldwide.^17,18^ Therefore, decreasing unnecessary antibiotic prescription is of high public health priority.

Similarly to other respiratory tract infections, patients infected with RSV are often exposed to high rates of unnecessary antibiotic therapy.^9,19–25^ Furthermore, several studies have tried to assess the clinical variables associated with antibiotic misuse in RSV patients.^9,20–23,26^ However, these studies mostly had sample sizes or limited available clinical covariates.

In this study, we examine antibiotic misuse in children <2 years of age hospitalized for RSV-bronchiolitis between 2008 and 2018 in an Israeli hospital. We study the patient-specific and clinical covariates that might lead to unnecessary prescription of antibiotics to patients infected with RSV. In addition, we characterize the amount and types of antibiotics unnecessarily prescribed.

## Methods

### Data sources

In this retrospective study, we extracted data from medical records of all children <2 years of age admitted to Hillel Yaffe Medical Center, Israel, during 2008-2018 for RSV-bronchiolitis. Initially, laboratory records of all children with positive RSV results were retrieved. RSV was ascertained by patients’ respiratory samples (nasopharyngeal swabs or aspirates), tested for the presence of RSV antigen.^25^ RSV-bronchiolitis was determined based on both clinical symptoms, diagnosis and laboratory results. Information was collected from medical records on demographics (age, sex), clinical symptoms (e.g., fever, dyspnea, etc.), performing chest x-ray and x-ray findings, laboratory tests (cultures, and their results, complete blood count, etc.), treatment modalities and prior emergency room visits/ hospitalizations.

Children with evidence of bacterial pneumonia (based on x-ray findings as determined by a qualified radiologist and senior pediatrician), bacterial growth in urine and blood were considered as bacterial infections. Further ascertainment of the lack of a bacterial infection was determined by a manual examination of each of the records by a senior pediatrician, based on laboratory findings of leukocytes, neutrophils and c-reactive protein. All the records determined to have bacterial infections (n=212) were excluded from further analyses. Our cohort thus included a total of 1015 patients determined to have RSV-bronchiolitis without evidence of bacterial coinfection. Individuals for whom antibiotic treatment was missing (n=12) were also excluded, leaving 1003 patients in our cohort.

Unnecessary antibiotic treatment (misuse), the dependent variable, was defined as antibiotic prescription during hospitalizations of children with RSV-bronchiolitis who lacked evidence of bacterial coinfection. The independent variables included the following 11 binary variables: demographics (*female, ethnicity* - Arabs or Jews), clinical factors (*background diseases, tachypnea at admission*), prior emergency room visits (*prior emergency room visit*), drawing of bacterial cultures during hospitalization (*blood culture taken, urine culture take, CSF culture taken*), admission during November-March (*hospitalization during cold season*), and treatment modalities (*steroids given at community* and *inhalation at hospital*).

Additionally, 5 continuous variables were used: the age of patients at admission (*age, weeks*); the percent of oxygen saturation as measured at admission (*% saturation at admission*); the patients’ temperature measured at hospital admission (*body temperature at admission)*; duration of the patients’ hospitalization, measured in days (*Hospitalization duration, days*) and the time elapsed from January 1, 2008, measured in years (*Years since 2008*).

All other variables used in the study, except for the types of antibiotics prescribed, had <3.5% missing values. The types of antibiotics prescribed were registered manually during the study period. Hence these data were available only for 280 of the 414 children who received unnecessary antibiotic treatment (67.6%).

### Statistical analysis

The uncertainty regarding the proportion of unnecessary antibiotic treatment was estimated using a binomial confidence interval. The data presented in Table 1 are based on a complete case analysis. The p-values presented in Table 1 are derived from t-tests for continuous variables and Fisher’s exact test for binary variables.

**Table 1:** Patient characteristics, stratified by (unnecessary) antibiotic treatment in the hospital. Mean (standard deviation) are presented for continuous variables; total number (%) are presented for binary variables. AB, antibiotics prescribed; CSF, cerebrospinal fluid.

|  | **No AB** | **AB** | **p-value** |
| --- | --- | --- | --- |
| **n** | 668 | 335 |  |
| **Age, weeks, mean (SD)** | 20.7 (18.6) | 27 (23.6) | <0.001 |
| **Female, n (%)** | 264 (39.5) | 154 (46.0) | 0.059 |
| **Prior emergency room visit, n (%)** | 24 ( 3.6) | 30 ( 9.0) | 0.001 |
| **% Saturation at admission, mean (SD)** | 96.9 (2.7) | 95.3 (5.3) | <0.001 |
| **Blood culture taken, n (%)** | 284 (42.8) | 233 (70.8) | <0.001 |
| **Urine culture taken, n (%)** | 50 ( 7.5) | 79 (23.9) | <0.001 |
| **CSF culture taken, n (%)** | 8 ( 1.2) | 41 (12.2) | <0.001 |
| **Tachypnea at admission, n (%)** | 443 (66.3) | 240 (71.9) | 0.089 |
| **Hospitalization during cold season, n (%)** | 640 (95.8) | 321 (95.8) | 1.00 |
| **Steroids given at community, n (%)** | 131 (20.3) | 54 (16.6) | 0.195 |
| **Background diseases, n (%)** | 51 (7.6) | 44 (13.1) | 0.007 |
| **Inhalation at hospital, n (%)** | 560 (83.8) | 287 (86.2) | 0.38 |
| **Body temperature at admission, mean (SD)** | 37.6 (0.8) | 38.2 (1) | <0.001 |
| **Arab ethnicity, n (%)** | 303 (45.4) | 143 (42.7) | 0.46 |
| **Hospitalization duration, days, mean (SD)** | 3.15 (2.47) | 4.60 (2.90) | <0.001 |
| **Years since 2008, mean (SD)** | 6 (2.8) | 6.3 (2.7) | 0.152 |

We modelled the relationship between the covariates and antibiotic administration (coded as a binary variable with 1=antibiotics administered and 0 otherwise). All missing values were imputed using a random forest algorithm from the *randomForest* R package^27^ before input into the logistic regression model.

The presented logistic regression model was selected by first including all variables that were deemed clinically relevant for the patient decision and were available to the physician before antibiotic prescription was performed. Then, backwards elimination was performed using the Akaike information criterion (AIC) to reach the final model.

All continuous variables in the final logistic regression model were also modeled in a varying coefficient framework, using generalized additive models with the mgcv *R* package.^28^ These resulted in approximately linear relationships, strengthening the rationale to keep these variables continuous.

### Ethical statement

The study was approved by the Institutional Review Board (Helsinki) Committee of Hillel Yaffe Medical Center. Since this was a retrospective study, using archived medical records, an exemption from informed consent was granted by the Helsinki Committee.

## Results

Overall, 1227 children aged <2 years were hospitalized for RSV-bronchiolitis during the study period. These were further subsetted to include only patients who were not additionally infected by a bacterial pathogen during their hospitalization, and for whom antibiotic administration was recorded, retaining 1003 patients. Of these patients, 335 received unnecessary antibiotic treatment. Hence, we estimate the proportion of unnecessary antibiotic treatment in our study population, which may include patients not appearing in our data, as 33.4% (95% CI 30.5%-36.4%).

Strong clinical suspicion of a bacterial infection should incentivise physicians to draw bacterial cultures, potentially followed by empirical antibiotic therapy. Indeed, bacterial cultures (either blood, urine, cerebrospinal fluid (CSF), or any combination of those) were drawn from 525 (52.4%) patients. However, antibiotics were nonetheless unnecessarily administered to 88 (19%) of patients for whom no bacterial cultures were drawn.

Based on these high rates of antibiotic misuse, we sought to understand what might be the reasons for unnecessary antibiotic administration to RSV patients. First, we present potentially relevant patient characteristics in Table 1, stratified by antibiotic administration within the hospital.

Next, we modeled the relationship between patients’ characteristics and antibiotic misuse through a logistic regression model. Importantly, some of the variables presented in Table 1 are not necessarily available to physicians before they decide whether to administer antibiotic treatment. Accordingly, we excluded the two variables from the logistic regression models: inhalation at the hospital and the hospitalization duration.Whereas the former was similar between the treated and untreated groups, the latter was not (Table 1).

After a model selection process, we remained with 9 variables, all serving as predictors of unnecessary antibiotic treatment among patients infected solely by RSV (Table 2). The model had a good fit to the data, attaining a C-statistic (equivalently, the area under the receiver-operator curve, AUC) of 0.785.

**Table 2:** Coefficients (exponentiated) of the multiple logistic regression model with antibiotic misuse as the dependent variable (prescribed antibiotics coded as one). Abbreviations: CSF, cerebrospinal fluid; CI, confidence interval; OR, odds ratio. Coefficients for continuous variables (age, saturation and body temperature) indicate the OR for an increase in one unit of the variable.

| *Predictors* | *Odds Ratio* | *95% CI* | *p-value* |
| --- | --- | --- | --- |
| **Female** | 1.41 | 1.04 – 1.92 | 0.029 |
| **Age, weeks** | 1.01 | 1.01 – 1.02 | 0.001 |
| **Prior emergency room visit** | 3.95 | 2.08 – 7.53 | <0.001 |
| **% Saturation at admission, continuous** | 0.88 | 0.84 – 0.93 | <0.001 |
| **Blood culture taken** | 2.46 | 1.75 – 3.48 | <0.001 |
| **Urine culture taken** | 2.89 | 1.76 – 4.75 | <0.001 |
| **CSF culture taken** | 6.35 | 2.77 – 16.08 | <0.001 |
| **Tachypnea at admission** | 1.75 | 1.24 – 2.50 | 0.002 |
| **Body temperature at admission** | 1.55 | 1.29 – 1.86 | <0.001 |

As might be expected, drawing of any bacterial culture increased the likelihood of antibiotic treatment. Similarly, variables indicating a “sicker” child were also positively associated with antibiotic treatment: prior admission to the emergency room; tachypnea, high temperature and lower oxygen saturation at admission. In contrast, older children were less likely to receive antibiotic treatment, as were male patients, although notably the regression coefficient for the latter variable had the highest p-value (0.029) in our model.

Finally, we examined the distribution of the antibiotics unnecessarily administered to the patients. Of the 335 children who received unnecessary antibiotic treatment, information about the antibiotic type was available only for 216 children (64.5%, see Methods). These summed to a total of 283 prescribed antibiotics. The distribution of the number of antibiotics prescribed was 56.1% (n=159), 17% (n=48), 2.8% (n=8) and 0.4% (n=1) for one, two, three and four antibiotics per patient, respectively. The most common prescriptions of at least two antibiotics were ampicillin and gentamicin for 13% of children (n=28), and amoxicillin and cefuroxime for 6.5% of children (n=14), with the rest of multiple prescriptions given for less than 2.5% (n=5) of children. We aggregated first- and third-generation cephalosporins (cefazolin and cefalexin, ceftriaxone and cefotaxime, respectively) and penicillins (amoxicillin and piperacillin) due to their similar clinical use and mechanisms of action. Under these groups we found that the most commonly prescribed antibiotics were penicillines (39.2%), cefuroxime (26.1%) 3rd generation cephalosporins (13.1%) gentamicin (10.2%) and azithromycin (8.5%), amoxicillin/clavulanic (1.8%) acid; 1st gen ceph (0.7%), and piperacillin/tazobactam (0.4%) (Figure 1).

**Figure 1:**
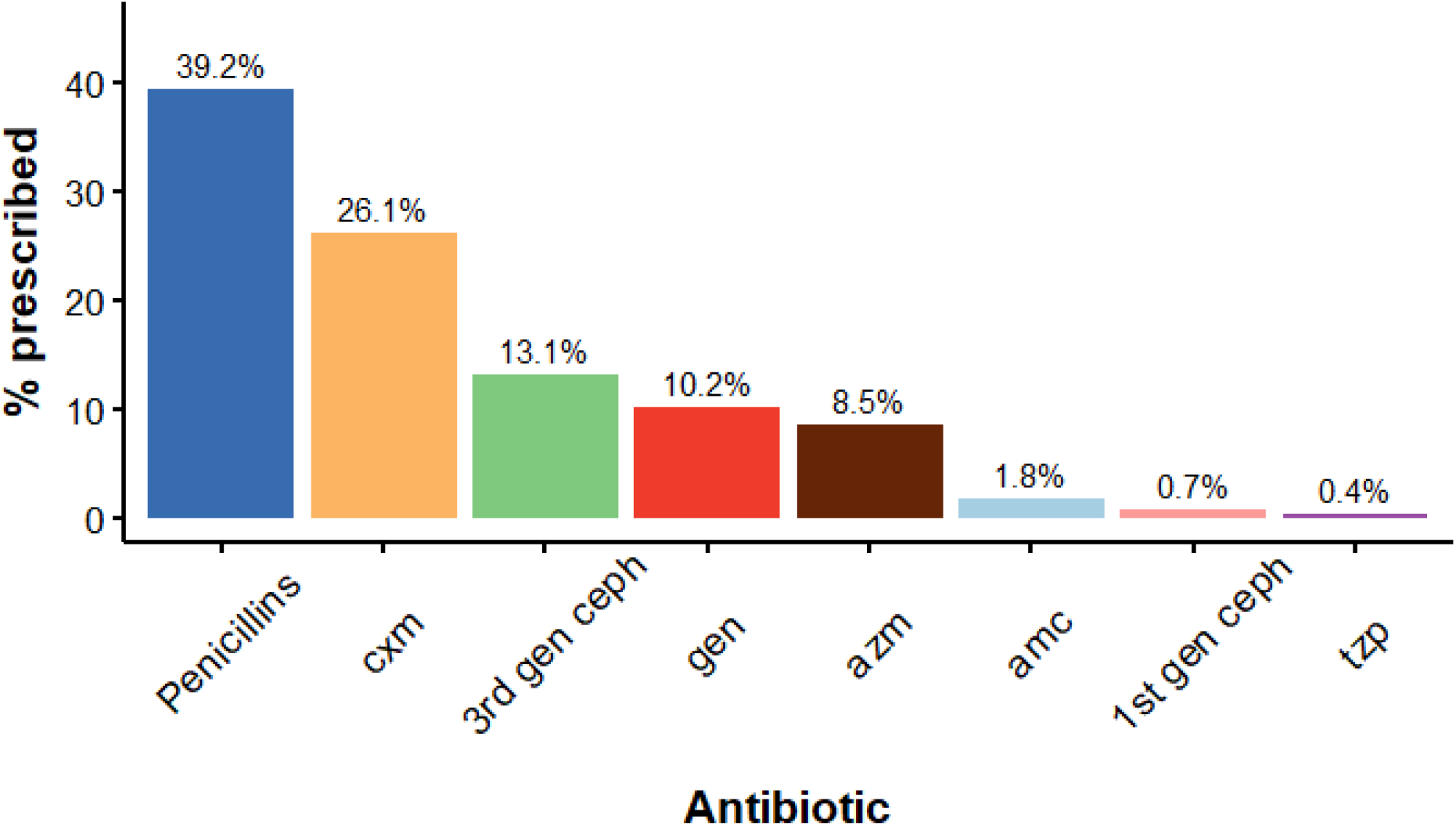
Distribution of antibiotics unnecessarily prescribed to RSV patients. Data were available for 213 patients, for whom n=283 antibiotics were prescribed. Abbreviations: Penicillins, amoxicillin and piperacillin; cxm, cefuroxime; 3rd gen ceph, ceftriaxone and cefotaxime; gen, gentamicin; azm, azithromycin; amc, amoxicillin/clavulanic acid; 1st gen ceph, cefazolin and cefalexin; tzp, piperacillin/tazobactam.

## Discussion

We investigated the extent of and predictors for antibiotic misuse in children with RSV-bronchiolitis. We found that of all children infected with RSV, 27.3% were unnecessarily treated with antibiotics. When examining only children ascertained not to have a bacterial coinfection, unnecessary antibiotic treatment increased to 33.4%. These numbers are in the range of estimates observed in other studies, although between-study variability is high.^9,19–25^

Using a multivariate logistic model, we found predictors that explain some of the physicians’ decision making process leading to administration of unnecessary antibiotic therapy. Drawing of blood, urine or CSF cultures had substantial associations with unnecessary antibiotic therapy.

This is in line with a strong suspicion of a bacterial infection, and a consequent administration of empiric antibiotic therapy. However, even in the absence of bacterial cultures, 19% of children with RSV-bronchiolitis still received unnecessary antibiotic treatment.

Other predictors found associated with increased antibiotic misuse were indicative of a severe patient status: lower oxygen saturation, higher temperature, tachypnea and prior visit to the emergency room. Accordingly, previous qualitative studies have found that physicians tend to erroneously prescribe antibiotics to patients who seem at high risk for adverse outcomes.^10,11^ Similar patterns were identified in previous observational studies examining antibiotic misuse for patients with viral respiratory infections.^22,26^

Interestingly, we identified that female sex is an influential predictor for unnecessary antibiotic treatment. Higher rates of antibiotic overprescription were previously reported for adult women, even when correcting for prevalence of conditions necessitating antibiotic therapy (e.g. urinary tract infections).^29,30^ Hence, the observed association could potentially result from the same reasons leading to unnecessary antibiotic prescription for adult females, such as over-precaution. On the other hand, we can not rule out residual confounding effects or a spurious association, as this variable has the highest p-value in our model (0.029).

Surprisingly, we found that older ages, which might suggest a more robust health status, were positively associated with unnecessary antibiotic treatment. Nonetheless, a similar pattern was observed in a recents studies. Children of older ages were shown to have increased likelihood of antibiotic treatment for viral respiratory tract infections^26^ and for febrile illness.^31^ Hence our findings support the notion that physicians might consider older children as more likely to have bacterial respiratory tract infections.

Additionally, we found that the most common (>2%) unnecessarily prescribed antibiotics were penicillines (39.2%), cefuroxime (26.1%) 3rd generation cephalosporins (13.1%) gentamicin (10.2%) and azithromycin (8.5%). With the exception of gentamicin, all the above listed antibiotics are usually employed to treat bacterial respiratory infections.^32^ Hence, most of the unnecessary treatments were indeed consistent with diagnoses of respiratory tract infections, and can be potentially reduced by means distinguishing between the bacterial and viral infections. The prescription of gentamicin, however, is consistent with a suspicion of a urinary or skin infection. In these cases, the misdiagnosis was in regards to both the causative pathogen and the site of infection and could have been probably more easily avoided.

Our study has several notable strengths. We obtained a large sample size, yielding high statistical power; the hospitalized children had laboratory confirmed RSV infections, reducing the potential for misclassification bias; we had access to detailed medical records, allowing us to examine different relevant variables; and importantly, our data were obtained from a single medical center, over a decade. Thus our study could provide robust estimates and minimize variation that might arise by including multiple centers.

The main limitation of our study is that reliance on a single center might restrict the generalizability of our estimate of antibiotic misuse. Nonetheless, the study hospital is a general governmental facility which serves various population sub-groups. Importantly, we had a notable representation of the largest and second-largest ethnic subpopulations in Israel - Jews and Arabs (Table 1). These represent populations living in both urban and rural settings, from various socioeconomic backgrounds. Hence, our findings should be generalizable to additional populations in high-income countries and those with western-like medical care systems and universal health care, as is the case in Israel.

Unnecessary antibiotic prescription is mainly responsible for adverse consequences in longer time frames, and often on a population level rather than for a specific patient. Hence, reducing unnecessary antibiotic prescription should be accomplished by promoting protocols and guidelines which also take into account the deleterious effects of antibiotic misuse. A salient example for implementations of such principles is antibiotic stewardship programs, which have shown substantial effectiveness in reducing antibiotic misuse in patients with RSV infections.

^33,34^ Another promising direction for reduction of antibiotic misuse are decision support systems. These systems are based on statistical algorithms, which can help provide unbiased probabilities for correct antibiotic treatment based on patients’ electronic medical files. ^35–38^ However, raw probabilistic outputs of such systems may still not suffice to reduce unnecessary antibiotic prescriptions, due to their interpretation by physicians. ^39,40^ Nonetheless, as we have previously shown, discretizing outputs of such systems, e.g. into ‘low’ and ‘high’ probability of antibiotic treatment necessity, could help reduce this issue.^40^

To conclude, the findings of this study indicate that the high rates of unnecessary antibiotic therapy in RSV-bronchiolitis patients may be largely due to the physicians’ perception of disease severity. A better balance between prescribing and withholding antibiotics must be achieved to mitigate future rates of antibiotic resistance. Hopefully, the ongoing introduction of antimicrobial stewardship programs and decision support systems will facilitate this balance and decrease antibiotic misuse.

## Data Availability

Given IRB and legal restrictions, we cannot provide any access to individual-level data, which we obtained from archived medical records from a single medical center and were collected as part of patients' clinical care, and not for research purposes. Legal and ethical restrictions apply for secondary usage of these data in research. Readers may contact Uri Obolski (the corresponding author) for further information.

## References

(1) Pebody, R.; Moyes, J.; Hirve, S.; Campbell, H.; Jackson, S.; Moen, A.; Nair, H.; Simões, E. A.; Smith, P. G.; Wairagkar, N.; others. Approaches to Use the WHO Respiratory Syncytial Virus Surveillance Platform to Estimate Disease Burden. Influenza Other Respir. Viruses 2020, 14 (6), 615–621.

(2) Shi, T.; McAllister, D. A.; O’Brien, K. L.; Simoes, E. A.; Madhi, S. A.; Gessner, B. D.; Polack, F. P.; Balsells, E.; Acacio, S.; Aguayo, C.; others. Global, Regional, and National Disease Burden Estimates of Acute Lower Respiratory Infections Due to Respiratory Syncytial Virus in Young Children in 2015: A Systematic Review and Modelling Study. The Lancet 2017, 390 (10098), 946–958.

(3) Obando-Pacheco, P.; Justicia-Grande, A. J.; Rivero-Calle, I.; RodrÍguez-Tenreiro, C.; Sly, P.; Ramilo, O.; MejÍas, A.; Baraldi, E.; Papadopoulos, N. G.; Nair, H.; others. Respiratory Syncytial Virus Seasonality: A Global Overview. J. Infect. Dis. 2018, 217 (9), 1356–1364.

(4) Kassem, E.; Na’amnih, W.; Bdair-Amsha, A.; Zahalkah, H.; Muhsen, K. Comparisons between Ethnic Groups in Hospitalizations for Respiratory Syncytial Virus Bronchiolitis in Israel. Plos One 2019, 14 (4), e0214197.

(5) Glatman-Freedman, A.; Kaufman, Z.; Applbaum, Y.; Dichtiar, R.; Steiman, A.; Gordon, E.-S.; Keinan-Boker, L.; Shohat, T.; Haklai, Z. Respiratory Syncytial Virus Hospitalization Burden: A Nation-Wide Population-Based Analysis, 2000-2017. J. Infect. 2020, 81 (2), 297–303.

(6) Greenberg, D.; Dagan, R.; Shany, E.; Ben-Shimol, S.; Givon-Lavi, N. Incidence of Respiratory Syncytial Virus Bronchiolitis in Hospitalized Infants Born at 33–36 Weeks of Gestational Age Compared with Those Born at Term: A Retrospective Cohort Study. Clin. Microbiol. Infect. 2020, 26 (2), 256–e1.

(7) Dagan, R.; Landau, D.; Haikin, H.; Tal, A. Hospitalization of Jewish and Bedouin Infants in Southern Israel for Bronchiolitis Caused by Respiratory Syncytial Virus. Pediatr. Infect. Dis. J. 1993, 12 (5), 381–386.

(8) Tonkin-Crine, S.; Yardley, L.; Little, P. Antibiotic Prescribing for Acute Respiratory Tract Infections in Primary Care: A Systematic Review and Meta-Ethnography. J. Antimicrob. Chemother. 2011, 66 (10), 2215–2223.

(9) van Houten, C. B.; Cohen, A.; Engelhard, D.; Hays, J. P.; Karlsson, R.; Moore, E.; Fernández, D.; Kreisberg, R.; Collins, L. V.; de Waal, W.; others. Antibiotic Misuse in Respiratory Tract Infections in Children and Adults—a Prospective, Multicentre Study (TAILORED Treatment). Eur. J. Clin. Microbiol. Infect. Dis. 2019, 38 (3), 505–514.

(10) Rodrigues, A. T.; Roque, F.; Falcão, A.; Figueiras, A.; Herdeiro, M. T. Understanding Physician Antibiotic Prescribing Behaviour: A Systematic Review of Qualitative Studies. Int. J. Antimicrob. Agents 2013, 41 (3), 203–212.

(11) Boiko, O.; Burgess, C.; Fox, R.; Ashworth, M.; Gulliford, M. C. Risks of Use and Non-Use of Antibiotics in Primary Care: Qualitative Study of Prescribers’ Views. BMJ Open 2020, 10 (10), e038851.

(12) Klein, E. Y.; Schueller, E.; Tseng, K. K.; Morgan, D. J.; Laxminarayan, R.; Nandi, A. The Impact of Influenza Vaccination on Antibiotic Use in the United States, 2010–2017. In Open forum infectious diseases; Oxford University Press US, 2020; Vol. 7, p ofaa223.

(13) Ryu, S.; Kim, S.; Kim, B. I.; Klein, E. Y.; Yoon, Y. K.; Chun, B. C. Temporal Relationship between Antibiotic Use and Respiratory Virus Activities in the Republic of Korea: A Time-Series Analysis. Antimicrob. Resist. Infect. Control 2018, 7 (1), 1–8.

(14) Alsan, M.; Morden, N.; Gottlieb, J. D.; Zhou, W.; Skinner, J. Antibiotic Use in Cold and Flu Season and Prescribing Quality: A Retrospective Cohort Study. Med. Care 2015, 53 (12), 1066.

(15) Costelloe, C.; Metcalfe, C.; Lovering, A.; Mant, D.; Hay, A. D. Effect of Antibiotic Prescribing in Primary Care on Antimicrobial Resistance in Individual Patients: Systematic Review and Meta-Analysis. Bmj 2010, 340.

(16) McFarland, L. Epidemiology, Risk Factors and Treatments for Antibiotic-Associated Diarrhea. Dig. Dis. 1998, 16 (5), 292–307.

(17) Pourmand, A.; Mazer-Amirshahi, M.; Jasani, G.; May, L. Emerging Trends in Antibiotic Resistance: Implications for Emergency Medicine. Am. J. Emerg. Med. 2017, 35 (8), 1172–1176.

(18) Aslam, B.; Wang, W.; Arshad, M. I.; Khurshid, M.; Muzammil, S.; Rasool, M. H.; Nisar, M. A.; Alvi, R. F.; Aslam, M. A.; Qamar, M. U.; others. Antibiotic Resistance: A Rundown of a Global Crisis. Infect. Drug Resist. 2018, 11, 1645.

(19) Bloomfield, P.; Dalton, D.; Karleka, A.; Kesson, A.; Duncan, G.; Isaacs, D. Bacteraemia and Antibiotic Use in Respiratory Syncytial Virus Infections. Arch. Dis. Child. 2004, 89 (4), 363–367.

(20) Samson, L. M.; Cooke, C.; MacDonald, N. E. ANALYSIS OF ANTIBIOTIC USE AND MISUSE IN CHILDREN HOSPITALIZED WITH RSV INFECTION.† 653. Pediatr. Res. 1996, 39 (4), 111–111.

(21) Wang, E. E.; Law, B. J.; Stephens, D.; others. Pediatric Investigators Collaborative Network on Infections in Canada (PICNIC) Prospective Study of Risk Factors and Outcomes in Patients Hospitalized with Respiratory Syncytial Viral Lower Respiratory Tract Infection. J. Pediatr. 1995, 126 (2), 212–219.

(22) Papan, C.; Willersinn, M.; Weiß, C.; Karremann, M.; Schroten, H.; Tenenbaum, T. Antibiotic Utilization in Hospitalized Children under 2 Years of Age with Influenza or Respiratory Syncytial Virus Infection–a Comparative, Retrospective Analysis. BMC Infect. Dis. 2020, 20 (1), 1–10.

(23) Akande, M. Y.; Fergie, J. E. Antibiotic Use and Length of Stay in Children Hospitalized with RSV Lower Respiratory Tract Infection (LRTI); Am Acad Pediatrics, 2017.

(24) Kadmon, G.; Feinstein, Y.; Lazar, I.; Nahum, E.; Sadot, E.; Adam, D.; Zamir, G.; Chodick, G.; Schiller, O. Variability of Care of Infants With Severe Respiratory Syncytial Virus Bronchiolitis: A Multicenter Study. Pediatr. Infect. Dis. J. 2020, 39 (9), 808–813.

(25) van Houten, C. B.; Naaktgeboren, C.; Buiteman, B. J.; van der Lee, M.; Klein, A.; Srugo, I.; Chistyakov, I.; de Waal, W.; Meijssen, C. B.; Meijers, P. W.; others. Antibiotic Overuse in Children with Respiratory Syncytial Virus Lower Respiratory Tract Infection. Pediatr. Infect. Dis. J. 2018, 37 (11), 1077–1081.

(26) Desai, N. M.; Sadlowski, J. L.; Mistry, R. D. Antibiotic Prescribing for Viral Respiratory Infections in the Pediatric Emergency Department and Urgent Care. Pediatr. Infect. Dis. J. 2020, 39 (5), 406–410.

(27) Liaw, A.; Wiener, M.; others. Classification and Regression by RandomForest. R News 2002, 2 (3), 18–22.

(28) Wood, S.; Wood, M. S. Package ‘Mgcv.’ R Package Version 2015, 1, 29.

(29) Schröder, W.; Sommer, H.; Gladstone, B. P.; Foschi, F.; Hellman, J.; Evengard, B.; Tacconelli, E. Gender Differences in Antibiotic Prescribing in the Community: A Systematic Review and Meta-Analysis. J. Antimicrob. Chemother. 2016, 71 (7), 1800–1806.

(30) Smith, D. R.; Dolk, F. C. K.; Smieszek, T.; Robotham, J. V.; Pouwels, K. B. Understanding the Gender Gap in Antibiotic Prescribing: A Cross-Sectional Analysis of English Primary Care. BMJ Open 2018, 8 (2), e020203.

(31) Van de Maat, J.; Van De Voort, E.; Mintegi, S.; Gervaix, A.; Nieboer, D.; Moll, H.; Oostenbrink, R.; Moll, H. A.; van Veen, M.; Noordzij, J. G.; others. Antibiotic Prescription for Febrile Children in European Emergency Departments: A Cross-Sectional, Observational Study. Lancet Infect. Dis. 2019, 19 (4), 382–391.

(32) Centers for Disease Control and Prevention. Pediatric Outpatient Treatment Recommendations. https://www.cdc.gov/antibiotic-use/clinicians/pediatric-treatment-rec.html.

(33) Quintos-Alagheband, M. L.; Noyola, E.; Makvana, S.; El-Chaar, G.; Wang, S.; Calixte, R.; Krilov, L. R. Reducing Antibiotic Use in Respiratory Syncytial Virus—A Quality Improvement Approach to Antimicrobial Stewardship. Pediatr. Qual. Saf. 2017, 2 (6).

(34) Kalil, J.; Bowes, J.; Reddy, D.; Barrowman, N.; Le Saux, N. Pediatric Inpatient Antimicrobial Stewardship Program Safely Reduces Antibiotic Use in Patients with Bronchiolitis Caused by Respiratory Syncytial Virus: A Retrospective Chart Review. Pediatr. Qual. Saf. 2019, 4 (5).

(35) Yelin, I.; Snitser, O.; Novich, G.; Katz, R.; Tal, O.; Parizade, M.; Chodick, G.; Koren, G.; Shalev, V.; Kishony, R. Personal Clinical History Predicts Antibiotic Resistance of Urinary Tract Infections. Nat. Med. 2019, 25 (7), 1143–1152.

(36) Lewin-Epstein, O.; Baruch, S.; Hadany, L.; Stein, G. Y.; Obolski, U. Predicting Antibiotic Resistance in Hospitalized Patients by Applying Machine Learning to Electronic Medical Records. Clin. Infect. Dis. 2021, 72 (11), e848–e855.

(37) Cherny, S. S.; Nevo, D.; Baraz, A.; Baruch, S.; Lewin-Epstein, O.; Stein, G. Y.; Obolski, U. Revealing Antibiotic Cross-Resistance Patterns in Hospitalized Patients through Bayesian Network Modelling. J. Antimicrob. Chemother. 2021, 76 (1), 239–248.

(38) Leibovici, L.; Paul, M.; Nielsen, A. D.; Tacconelli, E.; Andreassen, S. The TREAT Project: Decision Support and Prediction Using Causal Probabilistic Networks. Int. J. Antimicrob. Agents 2007, 30, 93–102.

(39) Poses, R. M.; Cebul, R. D.; Wigton, R. S. You Can Lead a Horse to Water-Improving Physicians’ Knowledge of Probabilities May Not Affect Their Decisions. Med. Decis. Making 1995, 15 (1), 65–75.

(40) Diamant, M.; Baruch, S.; Kassem, E.; Muhsen, K.; Samet, D.; Leshno, M.; Obolski, U. A Game Theoretic Approach Reveals That Discretizing Clinical Information Can Reduce Antibiotic Misuse. Nat. Commun. 2021, 12 (1), 1–13.

